# Determinants of infant formula feeding in Debre Berhan city: A community-based cross-sectional study

**DOI:** 10.1101/2024.05.21.24307601

**Authors:** Fitsum Zekarias, Alemtsehaye Gashu, Agmasie Damtew, Michael Amera, Besufikad Mulugeta, Solomon Hailemeskel

## Abstract

**Introduction:** Exclusively breastfed infants experience more rapid growth in the first 6 months than other infants. This is because exclusive breastfeeding offers infants a food source (breast milk) that is packed with essential nutrients, immune-boosting elements, and other biologically active compounds crucial for optimal growth and development. However, owing to the increasingly expanding market for breast milk substitutes, the duration of breastfeeding is declining worldwide.

**Objective:** To assess the determinants of infant formula feeding among mothers of infants aged 0-6 months in Debre Berhan city, 2023

**Methods and Materials:** Between the 10^th^ and 20^th^ of May 2023, a total of 656 mothers were selected by employing a multistage sampling technique. The data were collected using a semi-structured questionnaire, and the results are presented using frequencies, percentages, and graphs. In addition, bivariate and multivariable logistic regression models were employed to identify predictors of infant formula feeding. According to the multivariable logistic regression models, variables with a p value less than 0.05 were considered statistically significant predictors of infant formula feeding, and the adjusted odds ratio (AOR) was used to indicate the degree of association between the predictor variables and infant formula feeding.

**Results:** According to our study, 39.7% of the mothers in Debre Berhan city practiced infant formula feeding. In addition, mothers who were aged 25-34 years [AOR = 2.68, 95% CI: 1.26–5.70] or 35-45 years [AOR = 4.74, 95% CI: 1.86–12.1], primipara [AOR = 4.48, 95% CI: 2.69–7.45], had received antenatal care (ANC) [AOR = 2.26, 95% CI: 1.74–5.06], had delivered through the caesarian section (C/S) [AOR = 4.28, 95% CI: 2.12–8.65], not aware of the risks of infant formula feeding [AOR = 7.26, 95% CI: 4.09–12.85], or who had initiated breastfeeding after an hour of birth [AOR = 5.27, 95% CI: 3.14–8.85] were more likely to feed their babies infant formula.

**Conclusion:** Our findings suggest a high prevalence of infant formula feeding in Debre Berhan city, and the majority of the factors that lead to infant formula feeding are modifiable if proper interventions are implemented.

## Introduction

Exclusively breastfed infants experience more rapid growth in the first 6 months than other infants [1]. This is because exclusive breastfeeding offers infants a food source (breast milk) that is packed with essential nutrients, immune-boosting elements, and other biologically active compounds crucial for optimal growth and development [2,3]. However, owing to the continuously expanding market for breast milk substitutes, which grew by 41% between 2008–2013, the duration of breastfeeding is declining worldwide [4–6].

Infant formula is manufactured industrially to serve as an alternative to breast milk, and to produce different classes of infant formula, breast milk substitutes manufacturing companies primely use modified cow milk or soy protein [7,8]. Although infant formula is manufactured in compliance with *Codex Alimentarius* standards to meet the nutritional needs of infants for up to six months, it does not fully replace the ideal food for infants, which is breast milk [7,9].

This is because breast milk possesses unique characteristics and ingredients that are absent in infant formula. These include the ability to alter its composition to cater to the specific needs of a child, an array of antibodies and other bioactive compounds that aid in combating infections, a perfect blend of nutrients that meet a child’s nutritional needs, and hypoallergenic properties that minimize the risk of allergic reactions [8,10]. Moreover, breast milk is rich in prebiotic and probiotic compounds that are essential for maintaining a healthy gut microbiome in newborns [8,10].

In the global market, infant formula is available in powder, liquid, or ready-to-feed forms [8]. The powdered form of infant formula, which is the commonest form in Ethiopia, is not sterile, and serious infections in children are traced back to this form of infant formula, which is contaminated with deadly bacteria, such as *Enterobacter sakazakii* [11]. Likewise, a study in Vietnam revealed a high risk of hospitalization among formula-fed infants from infections, such as otitis media, pneumonia, and diarrhea [12,13]. Furthermore, soy-based infant formula contains *phytoestrogens*, which can reduce the reproductive capability of both boys and girls later in life [14].

In addition, early initiation of infant formula feeding can disrupt the natural breastfeeding process, as it prolongs the commencement of breastfeeding and can lead to unnecessary termination of breastfeeding [15,16]. Such disruptions prevent both infants and mothers from reaping the full benefits of optimal breastfeeding, which includes a lower risk of infectious and chronic diseases, protection against certain types of cancer, appropriate spacing between births, enhanced intelligence and academic performance, a robust bond between a mother and her child, and a reduced cost of child rearing [10,15,17–29].

Despite the many benefits of breastmilk, globally, 11% of infants aged 0 to 5 months received infant formula in 2018, and the majority (37%) of those children were born in Latin America and Caribbean countries [5]. The reasons why mothers choose infant formula over breast milk are complex. However, factors such as the aggressive promotion of breast milk substitutes, negative social and cultural views on breastfeeding, work-related pressures, and inability to support mothers during breastfeeding are believed to play significant roles [15].

Globally, despite the commendable effort expended by the World Health Organization (WHO), which notably led to the introduction of the International Code of Marketing of Breastmilk Substitutes in 1981, infant formula feeding persists in both developed and developing nations [4,6,30]. For instance, a recent study in the U.S. reported that 20% of breastfeeding babies in 62% of maternity facilities nationwide were fed formula during their hospital stay [31]. Similarly, 40% of newborn babies in Australia were supplemented with breast milk substitutes before the age of six months [32].

In addition, according to a review that analyzed data from 85 countries, 27.7% of infants in low- and middle-income countries consumed commercially available breastmilk substitutes between 2010 and 2019, and the highest and lowest percentages were recorded in Gabon (63.5%) and Burkina Faso (0.8%), respectively [6]. Similarly, in Eastern and Southern Africa, approximately 4% of infants received infant formula in 2018 [5].

In Ethiopia, to promote optimal breastfeeding and reduce the increasing reliance on breast milk substitutes, in 2016, the government implemented various measures, including the introduction of the *National Nutrition Program II; the National Guidelines on Adolescent, Maternal, Infant, and Young Child Nutrition; the Food Advertisement Directive 33/2016; and the Infant Formula and Follow-up Formula Directive No. 30/2016* [33–36]. Unfortunately, despite such efforts, the percentage of children under two years of age who have received infant formula has doubled since 2016 (from 1.4% in 2016 to 3.3% in 2019) [35,37]. In addition, 41% of infants in the country are not currently exclusively breastfed, and nearly 16.5% and 10% of children received prelacteal and mixed feed (formula and/or animal milk in addition to breast milk), respectively [35,37,38].

Furthermore, multiple studies conducted in Mettu, Offa, Jimma, Bishoftu, Addis Ababa, Dire Dawa, Mekelle, and Gondar reported varying rates of formula feeding, ranging from 7.8% to 68.8% [39–46]. These studies have also identified factors such as educational status, cesarean delivery, prelacteal feeding, delayed breastfeeding initiation, knowledge and attitudes toward formula feeding, maternal age, occupational status, ANC utilization, source of information about breast milk substitutes, parity, and mode of delivery as important predictors of formula feeding among mothers in Ethiopia [39–46].

In conclusion, despite ongoing international and local efforts to promote optimal breastfeeding, a significant number of infants in Ethiopia are not optimally breastfed. In addition, depending on socioeconomic status, health service availability, and utilization patterns, the prevalence of infant formula feeding varies significantly across the country. Therefore, to provide contextual insights into this problem and also solidify the existing knowledge regarding the dynamics of infant formula feed in Ethiopia, this study was conducted in Debre Berhan city, where no similar studies have been conducted before, to assess the determinants of infant formula feeding. In addition, the authors believe that the findings of this study will have paramount importance in delivering more targeted interventions to reverse the rising level of infant formula feeding in Ethiopia.

## Materials and Methods

### Study area and period

The study was conducted between the 10^th^ and 20^th^ of May 2023 in Debre Berhan city, which is located 130 km and 695 km from the capital city of Ethiopia (Addis Ababa) and the regional capital city of Bahir Dar, respectively. The city is under North Shoa Zona in the Amhara regional state of Ethiopia. According to the 2015 population projection, a total of 202,226 people inhabited the city, 106,388 of whom were females. To address the health needs of its inhabitants, the city houses two hospitals, eight health centers, and eighteen health posts.

### Populations

This study considered mothers of infants aged 0-6 months who had lived in the study area for at least six months as a source population, whereas mothers of infants aged 0-6 months who were living in the selected kebeles were considered a study population. In addition, mothers who had medical reasons for providing infant formula to their babies and who were physically and mentally unfit to provide information were excluded from the study.

### Sample size determination

A sufficient number of participants who needed to be included in the study to accurately estimate the prevalence of infant formula feeding in Debre Berhan city was calculated using a single population proportion formula under the assumptions of 28.4% infant formula feeding [39], 5% margin of error, and 95% CI. Furthermore, the same size was adjusted for possible design effects (multiplied by 2) and nonresponses (5% added), resulting in a final sample size of 656.

### Sampling procedure

In this study, a multistage sampling technique was employed to select the study participants. To explain further, initially, from a total of five sub-cities within the Debre Berhan city three were selected using simple random sampling technique. Similarly, within each sub-city, four kebeles were selected using a simple random sampling technique. Subsequently, the number of eligible mothers in the selected kebeles were obtained from the health extension worker (as recorded in the expanded program of immunization (EPI) registration book), and using this information as a guide, a proportional allocation of the sample was performed. Finally, by employing a systematic random sampling technique, the study participants were selected from the EPI record book. **(Figure 1)**

**Figure 1:**
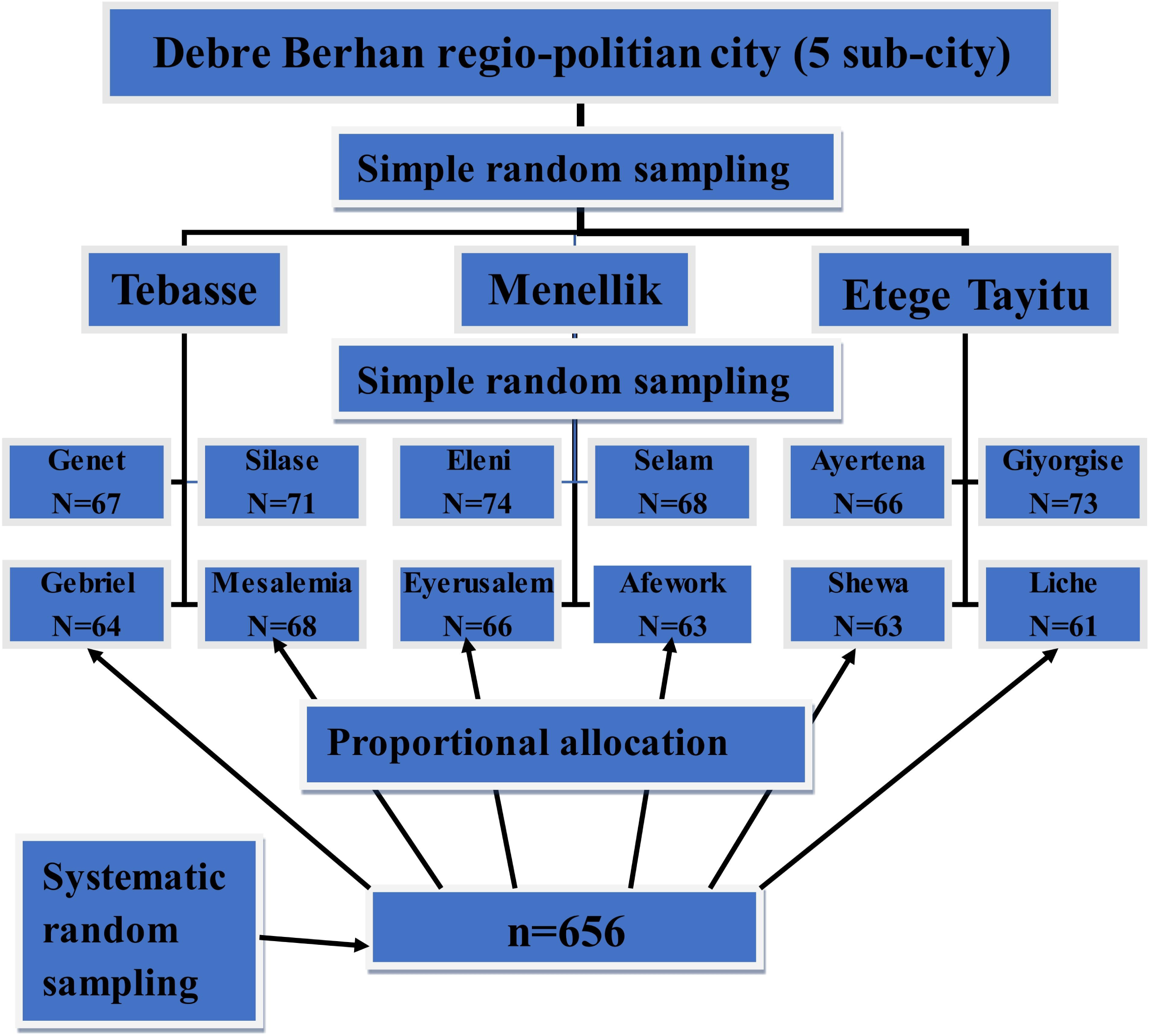
Schematic representation of the sampling procedure used to select mothers of infants aged 0-6 months in Debre Berhan city, North Showa Zone, Ethiopia, 2023.

### Data collection tool, technique, and quality control

The data were collected using a meticulously designed semi-structured questionnaire, which was designed based on insights gained during the review of prior studies with objectives akin to the current study [39–46]. The questionnaire was comprehensive and included queries related to sociodemographic attributes, obstetric history, infant feeding practices, and maternal knowledge of breastfeeding.

To ensure the reliability and consistency of the questionnaire, it was initially drafted in English, subsequently translated into Amharic (the local language), and retranslated back into English. To further validate the questionnaire, a pretest was conducted on 5% of the sample in a neighboring town. Moreover, each questionnaire was thoroughly checked for completeness before data entry.

### Data Processing and Analysis

Upon completion of the interviews, the data were inserted into Epi-data version 4.6 and subsequently exported to the Statistical Package for Social Sciences (SPSS) version 25 for in-depth statistical analysis. Within SPSS, the model’s fitness and the absence of multicollinearity were assessed using the Hosmer–Lemeshow goodness of fit (0.82) and the variance inflation factor (VIF < 5), respectively.

Before further analysis, the outcome variable was categorized as ‘1’ representing infant formula feeding and ‘2’ representing exclusive breastfeeding, and providing infant formula as a supplement to or substitute for breast milk within the past 24 hours was used as an indicator of infant formula feeding [39]. Furthermore, to measure mothers’ knowledge of breastfeeding, a set of seven questions was posed. Mothers who correctly answered four or more of these questions were deemed to possess adequate knowledge of breastfeeding.

Thereafter, descriptive analysis was performed, and the results are presented using frequencies, percentages, and graphs. In addition, bivariate and multivariable logistic regression models were employed to identify predictors of infant formula feeding. During the bivariate logistic regression analysis, a p value score of less than 0.25 was used as a criterion for selecting the variables to be included in the multivariable regression model. Finally, in the multivariable logistic regression analysis, variables with a p value score of less than 0.05 were considered statistically significant predictors of infant formula feeding, and the AOR at 95% CI was used to indicate the degree of association between the predictor variables and infant formula feeding.

### Ethical consideration

Ethical approval was obtained from the ethical review board of the Asrat Woldeyes Health Science campus, Debre Berhan University, in 2023 (**protocol number: IRB-147**). Additionally, the city health office wrote a letter of cooperation, which was subsequently submitted to the responsible bodies. Furthermore, both verbal and written informed consent were obtained from the study participant before each interview. To maintain the anonymity of the study participants, all their personal information was kept confidential.

## Results

In this study, a total of 635 mothers aged between 18 and 45 years were interviewed resulting in a response rate of 96.7%.

### Sociodemographic characteristics

Almost all of the mothers (97.3%) were married, and most (93.4%) followed the Orthodox religion. Regarding their education, more than half (53.9%) had received a college degree or above. However, approximately 37% of the mothers were not employed. Among their children, a large number (51.3%) were males, and most (78.4%) were younger than two or older than four months of age. In addition, 54.2% of the mothers’ spouses had received a college education or above, and the majority (41%) also worked in private institutions. **(Table 1)**

**Table 1.** Sociodemographic characteristics of mothers of infants aged 0-6 months in Debre Berhan city, North Shewa Zone, Ethiopia, 2023.

| Variables | Categories | Frequency | Percentage |
| --- | --- | --- | --- |
| Sex of the current child | Male | 326 | 51.3 |
|  | Female | 309 | 48.7 |
| **Age of the current child in months** | < 2 | 249 | 39.2 |
|  | 2-3 | 137 | 21.6 |
|  | > 4 | 249 | 39.2 |
| **Age gap between the current and the previous child in years (n=371)** | < 2 | 152 | 40.9 |
|  | ≥ 2 | 219 | 59.1 |
| **Age of the mother in years** | 15-24 | 85 | 13.4 |
|  | 25-34 | 434 | 68.3 |
|  | 35-45 | 116 | 18.3 |
| **Religion of the mother** | Orthodox | 593 | 93.4 |
|  | Protestant | 16 | 2.5 |
|  | Muslim | 26 | 4.1 |
| **Marital status** | Married | 618 | 97.3 |
|  | Unmarried | 17 | 2.7 |
| **Educational status of the mother** | Unable to read and write | 44 | 6.9 |
|  | can read and write | 20 | 3.1 |
|  | Primary | 85 | 13.4 |
|  | Secondary | 144 | 22.7 |
|  | College and above | 342 | 53.9 |
| **Occupation of the mother** | Private employee | 158 | 24.9 |
|  | Gov't employee | 183 | 28.8 |
|  | Daily laborer | 15 | 2.4 |
|  | Marchant | 39 | 6.1 |
|  | Farmer | 5 | 0.8 |
|  | Housewife | 235 | 37 |
| **Educational status of the father** | Unable to read and write | 15 | 2.4 |
|  | Can read and write | 15 | 2.4 |
|  | Primary | 71 | 11.2 |
|  | Secondary | 190 | 29.9 |
|  | College and above | 344 | 54.2 |
| **Occupation of the father** | Private employee | 264 | 41.6 |
|  | Gov't employee | 200 | 31.5 |
|  | Daily laborer | 55 | 8.7 |
|  | Marchant | 63 | 9.9 |
|  | Farmer | 32 | 5.0 |
|  | Unemployed | 21 | 3.3 |
| **Residence** | Urban | 609 | 95.9 |
|  | Rural | 26 | 4.1 |

### Obstetric and infant feeding-related characteristics

A high proportion (91.0%) of the mothers had at least one ANC visit during their pregnancy. However, a noteworthy percentage (82.7%) of the mothers missed postnatal care (PNC) appointments. The majority of mothers (94.6%) opted for childbirth in public health facilities, while a minority (5.4%) chose private health institutions or home births. During birth, more than three-quarters (78.7%) had undergone spontaneous vaginal delivery (SVD), while approximately 18% chose C/S. Moreover, slightly more than half (60.6%) of the mothers had initiated breastfeeding within the first hour after their child’s birth. **(Table 2)**

**Table 2.**
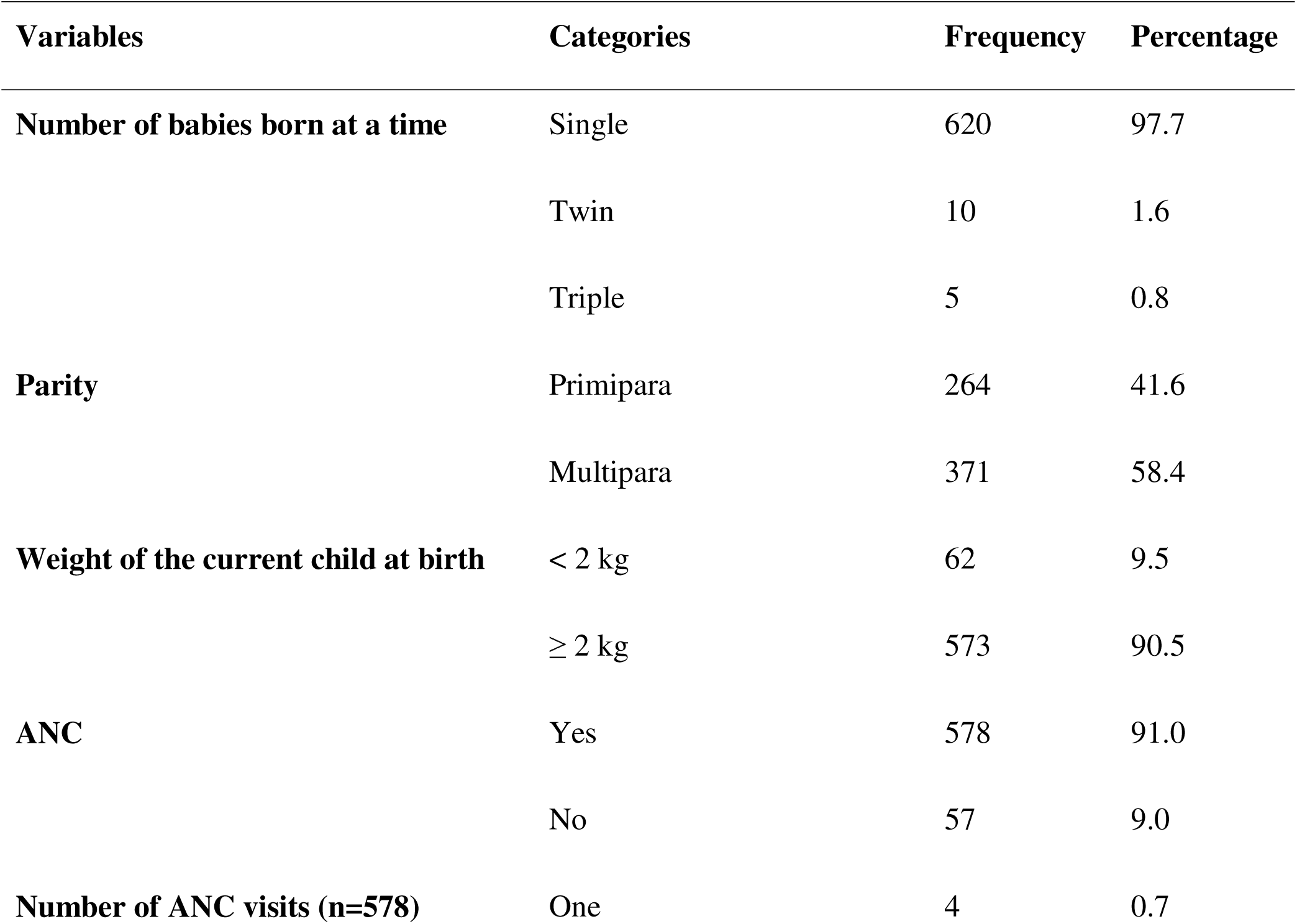
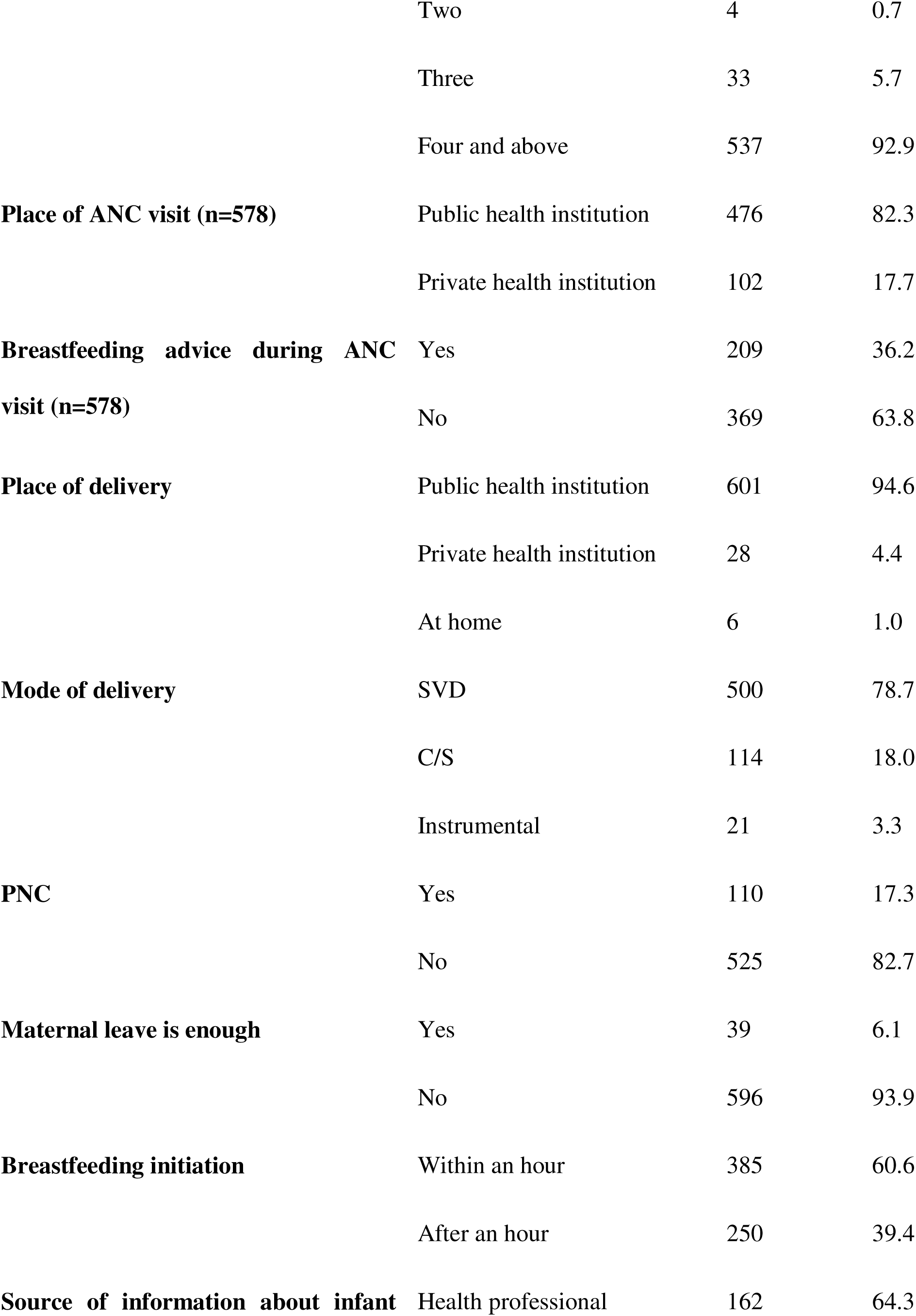
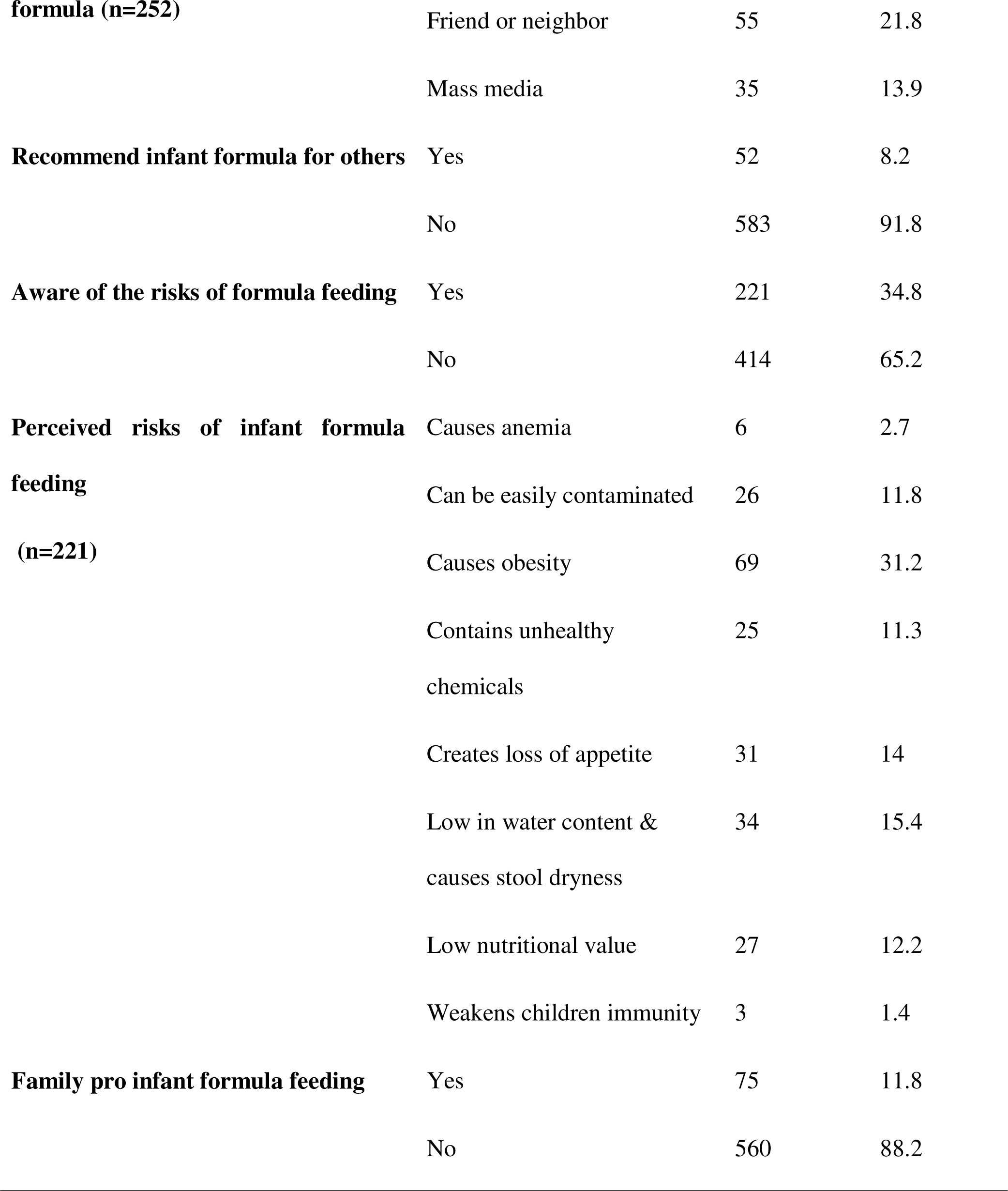
Obstetric and infant feeding practices of mothers of infants 0-6 months of age in Debre Berhan City, North Shewa Zone, Ethiopia, 2023.

### Mothers’ knowledge of breastfeeding

The majority of the mothers (77.3%) believed that breast milk alone was sufficient for the first six months. However, approximately one-quarter of them (22.7%) thought otherwise. In general, close to 27% of the mothers who participated in this study had poor knowledge of breastfeeding. **(Table 3**, **Figure 2)**

**Figure 2:**
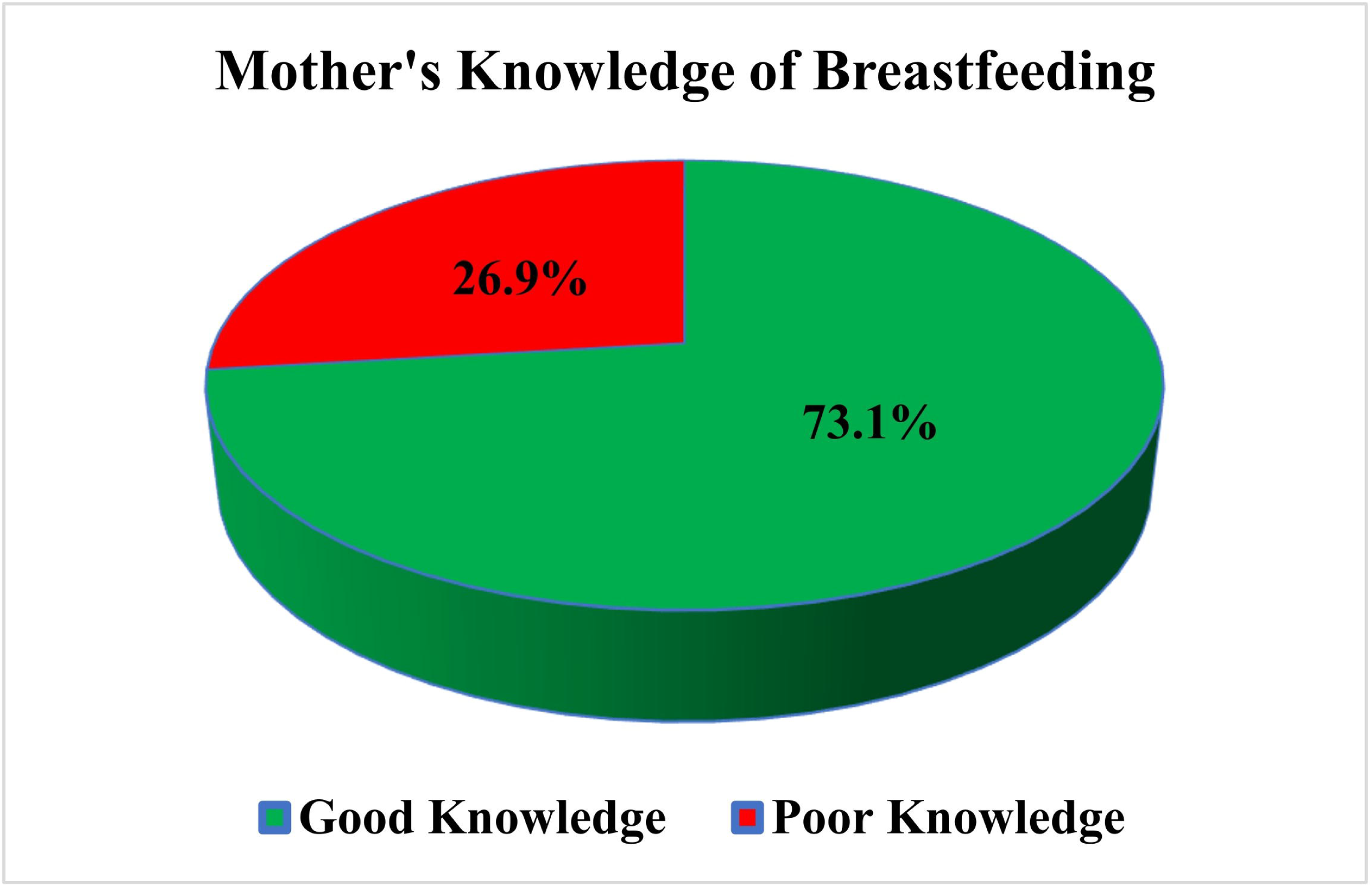
Breastfeeding knowledge of mothers of infants aged 0-6 months in Debre Berhan city, North Showa Zone, Ethiopia, 2023.

**Table 3.** Breastfeeding knowledge of mothers North Shewa Zone, Ethiopia, 2023 of infants aged 0-6 months in Debre Berhan city, North Shewa Zone, Ethiopia, 2023.

| Variables | Categories | Frequency | Percentage |
| --- | --- | --- | --- |
| **Breast milk alone is enough for the first six months** | True | 491 | 77.3 |
|  | False | 144 | 22.7 |
| **Children who are fed breast milk have a lower risk of contracting a disease** | True | 599 | 94.3 |
|  | False | 31 | 4.9 |
|  | I don't know | 5 | 0.8 |
| **Formula feeding incurs more cost than breastfeeding** | True | 571 | 89.9 |
|  | False | 31 | 4.9 |
|  | I don't know | 33 | 5.2 |
| **Breast milk contains all the nutrients necessary for optimal infant growth** | True | 586 | 92.3 |
|  | False | 16 | 2.5 |
|  | I don't know | 33 | 5.2 |
| **Breastfeeding promotes mother-to-child bond** | True | 565 | 89.0 |
|  | False | 26 | 4.1 |
|  | I don't know | 44 | 6.9 |
| **Breastfeeding delays the next pregnancy** | True | 322 | 50.7 |
|  | False | 153 | 24.1 |
|  | I don't know | 160 | 25.2 |
| **Breast milk has more benefits for children than formula milk** | True | 561 | 88.3 |
|  | False | 15 | 2.4 |
|  | I don't know | 59 | 9.3 |

### Magnitude of infant formula feeding

According to our study, 39.7% (95% CI: 35.6, 43.65) of the mothers in Debre Berhan city formula-fed their child, and the majority had done so because of insufficient breast milk. **(Figure 3**, **Figure 4)**

**Figure 3:**
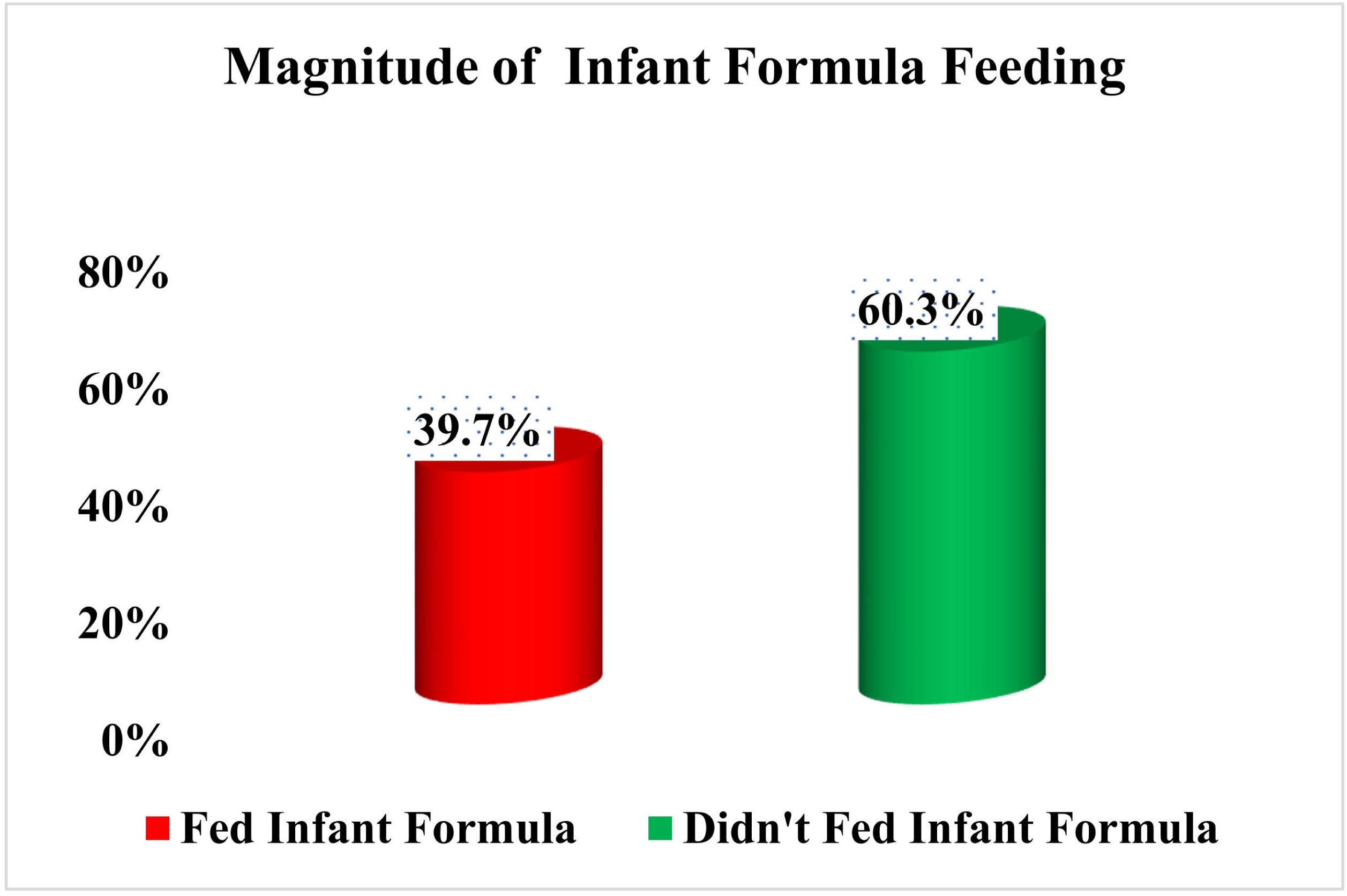
Magnitude of infant formula feeding among mothers of infants aged 0-6 months in Debre Berhan city, North Showa Zone, Ethiopia, 2023.

**Figure 4:**
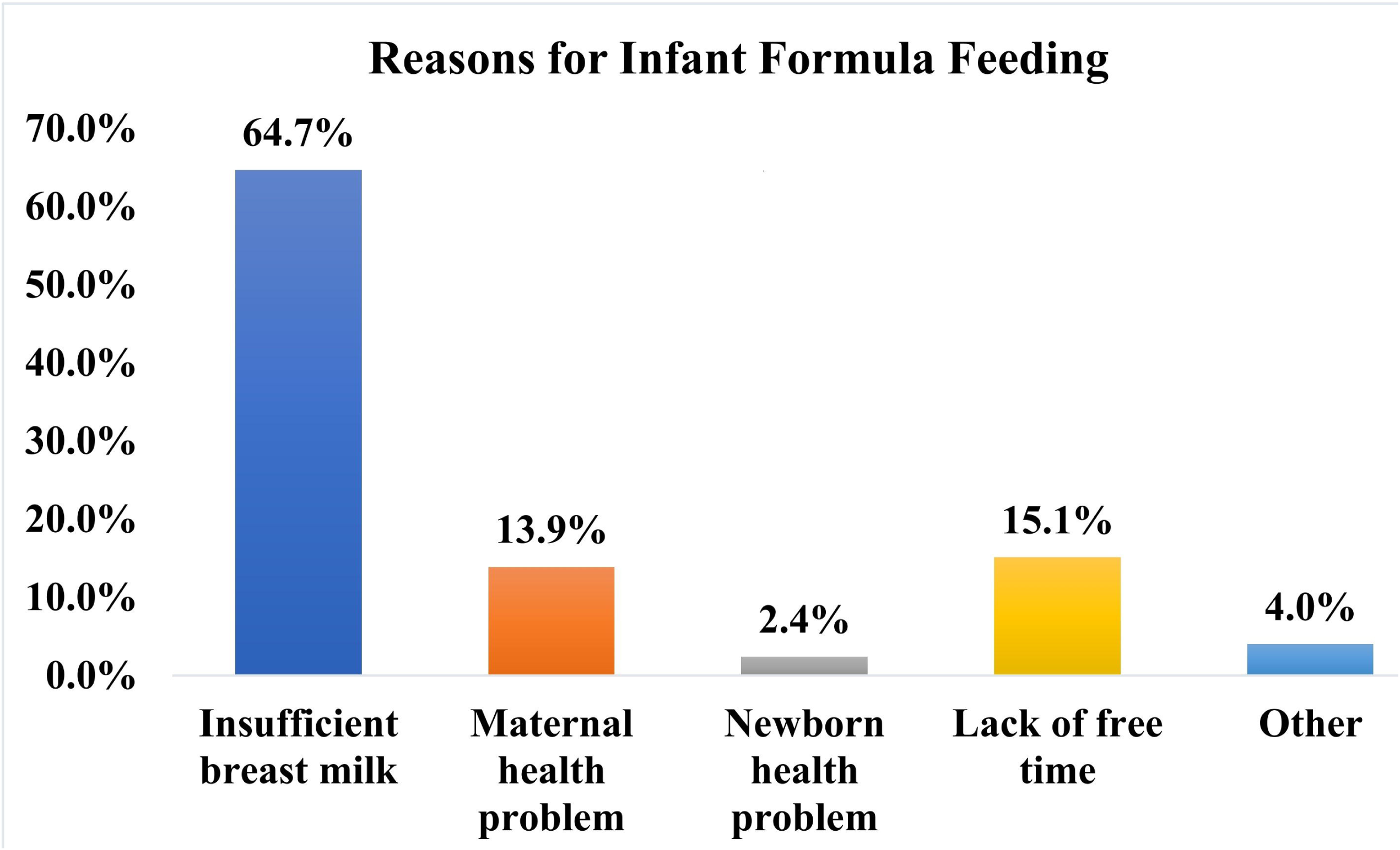
Reasons for infant formula feeding among mothers of infants aged 0-6 months in Debre Berhan city, North Showa Zone, Ethiopia, 2023.

### Factors associated with infant formula feeding

In this study, the associations of approximately thirty variables with infant formula feeding were investigated. According to the bivariate logistic regression analysis, twenty-five variables were significantly associated (p value <0.25) with infant formula feeding. However, after accounting for confounding variables in the multivariable logistic regression model, only six of the variables were found to be significantly associated with infant formula feeding (p value < 0.05).

Specifically, the odds of infant formula feeding were 2.68 and 4.74 times greater among mothers aged 25-34 years [AOR = 2.68, 95% CI: 1.26–5.70] and 35-45 years [AOR = 4.74, 95% CI: 1.86–12.1] respectively than among those aged 15-24 years. In addition, the odds of infant formula feeding were 5.27 times greater among mothers who initiated breastfeeding after an hour of delivery than among those who initiated breastfeeding within an hour of delivery [AOR = 5.27, 95% CI: 3.14– 8.85]. Furthermore, the likelihood of infant formula feeding was 7.26 times greater among mothers who were not aware of the risks of infant formula feeding than among those who were [AOR = 7.26, 95% CI: 4.09–12.85]. Moreover, mothers who had received ANC were 2.26 times more likely to provide infant formula than were those who had not received ANC [AOR = 2.26, 95% CI: 1.00– 5.06]. Additionally, the odds of infant formula feeding were 4.28 times greater among mothers who had delivered through C/S than among those who had delivered through SVD [AOR = 4.28, 95% CI: 2.12–8.65]. Finally, primiparous mothers were 4.48 times more likely to feed infant formula than multiparous mothers were [AOR = 4.48, 95% CI: 2.69–7.45]. **(Table 4)**

**Table 4.**
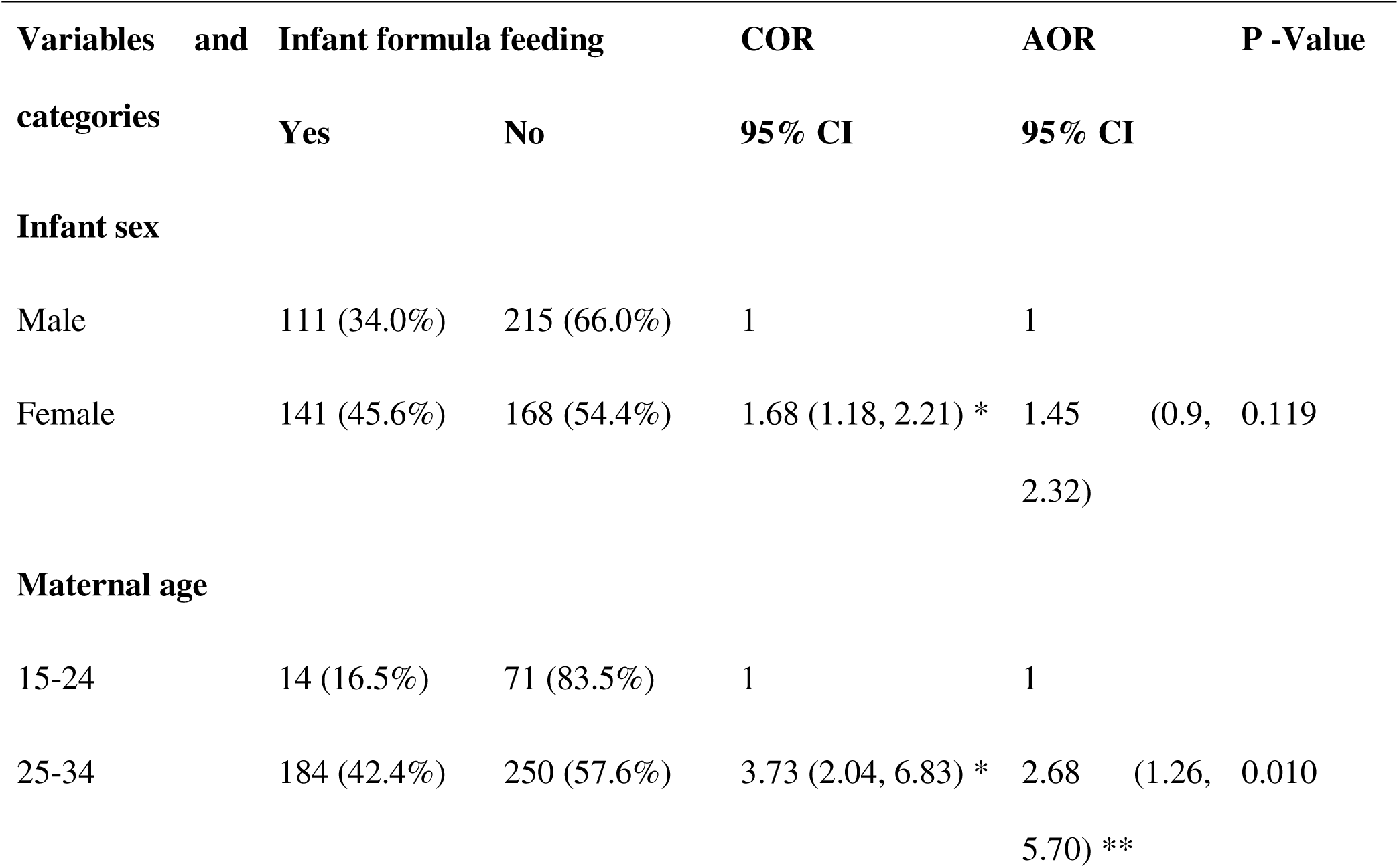
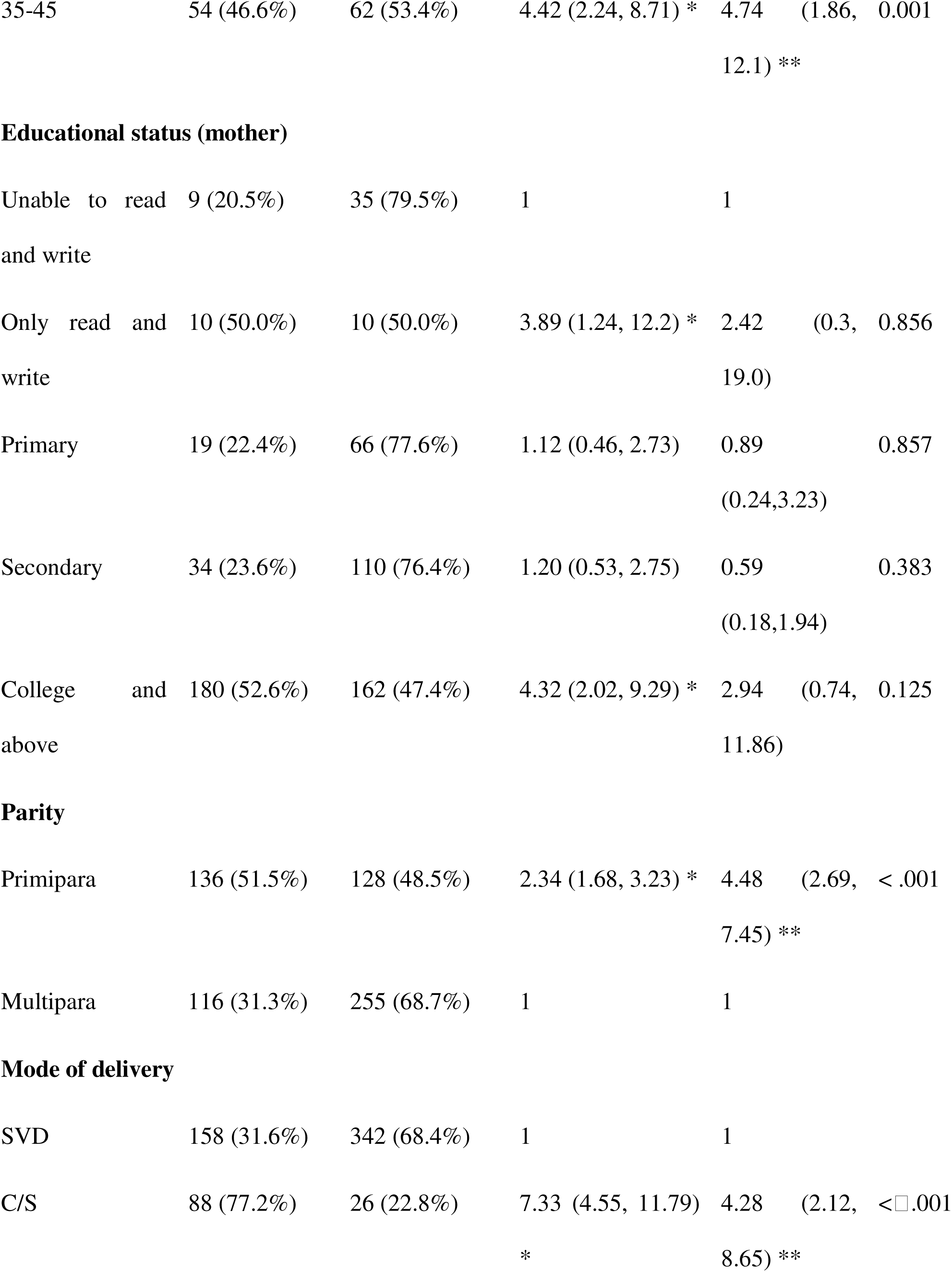
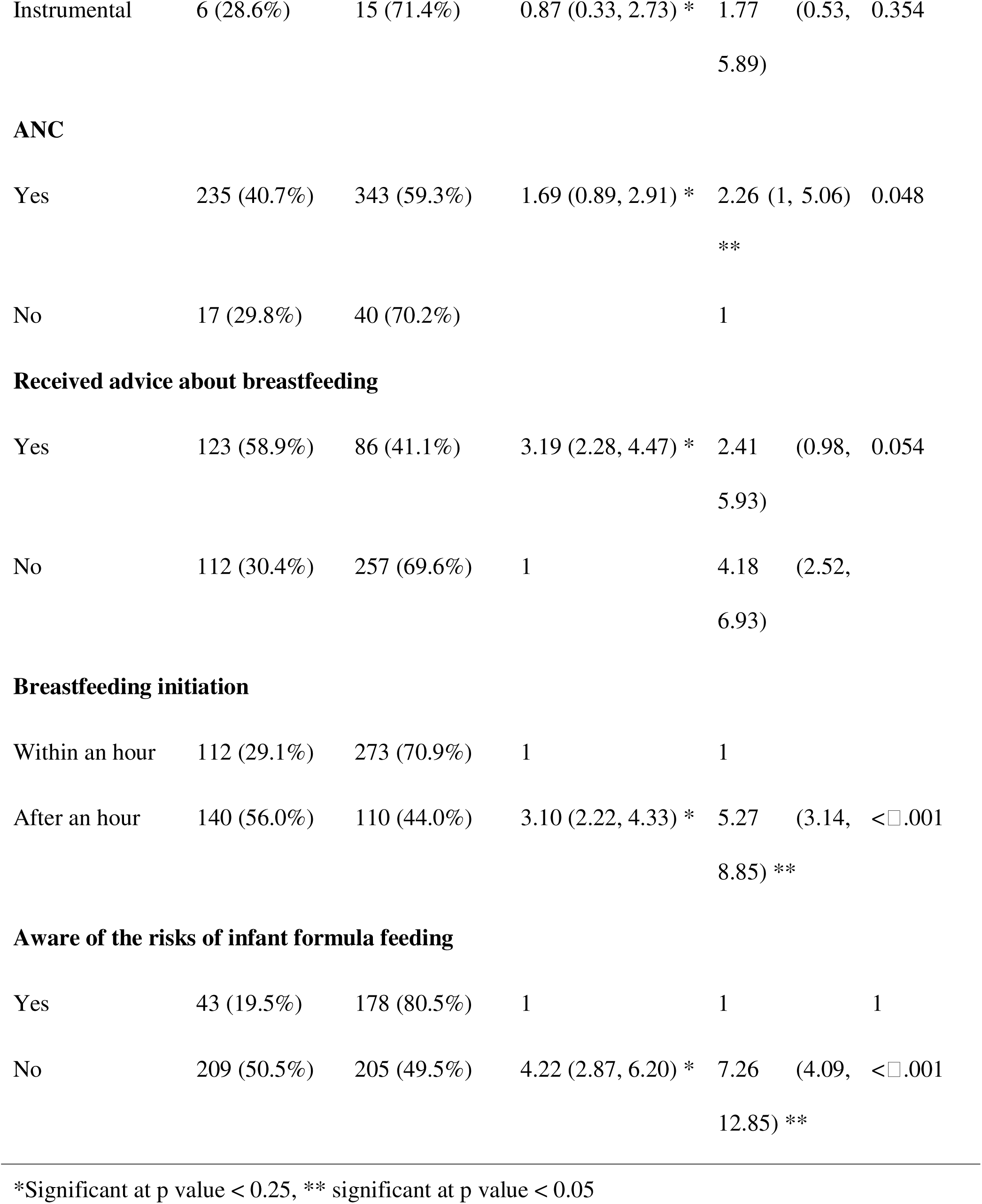
Factors associated with infant formula feeding among mothers of infants aged 0-6 months in Debre Berhan City, North Shewa Zone, Ethiopia, 2023.

## Discussion

In Debre Berhan city, nearly half of the mothers [39.7%, 95% CI: 35.6–43.65] practiced infant formula feeding. In addition, older mothers, mothers who initiated breastfeeding after an hour of birth, mothers who were not aware of the risk of infant formula feeding, mothers who had delivered through cesarean section, mothers who were primiparous, and mothers who had received ANC were more likely to practice infant formula feeding.

The percentage of infant formula-fed infants in Debre Berhan city was higher than that in Offa town (7.8%), Mettu town (28.4%), and Bahir Dar city (25%) [39,40,42]. In contrast, it was lower than the level found in Dire Dawa city (42.6%), Addis Ababa city (46.2%), and the Jimma Zone (47.2%) [41,44,45]. The misalignment between the findings of the current study and those of the studies presented above may stem from differences in the socioeconomic characteristics of the population as well as the area where the studies were carried out. For instance, a high prevalence of infant formula feeding was observed in major metropolitan cities, where a large number of distributors of infant formula as well as customers with deep pockets were located. In such places, the competitive environment by itself can force providers to flag their products using aggressive promotional tactics, such as the dissemination of misleading information about the potential use and risks of infant formula, to attract a greater number of potential buyers. As a result, clueless mothers, who had the means to purchase the products, might be tricked into buying and utilizing these products. However, such competitions are less likely to occur in small cities such as Debre Berhan. Therefore, differences in the level of infant formula feeding between such areas might be driven by other factors, such as variations in mothers’ educational level, amount of income, and level of healthcare utilization.

Our study also demonstrated that mothers who had not initiated breastfeeding within one hour of delivery were approximately five times more likely to practice infant formula feeding than were those who had initiated breastfeeding within one hour of delivery. This result is in agreement with the results of studies conducted in Addis Ababa city, Mettu town, and Bahir Dar city [39,41,42]. A possible justification might be that early initiation of breastfeeding fosters a strong bond between the mother and her baby, and such a strong relationship can lay the groundwork for mothers to spend more time with their babies and breastfeed them promptly. In addition, multiple studies have demonstrated that early initiation of breastfeeding significantly reduces the likelihood of prelacteal feeding in newborns, and one type of prelacteal feeding that is commonly practiced in hospitals is infant formula feeding [6,15]. Therefore, by promoting early breastfeeding, it is possible to significantly decrease the number of newborns who are receiving infant formula.

In addition to the abovementioned predictor, not being aware of the risks of infant formula feeding increased the odds of infant formula feeding by more than sevenfold. Similarly, a study in Mettu Town showed that the odds of infant formula feeding were two and a half times greater among mothers who lacked sufficient knowledge about infant formula than among those who had sufficient knowledge about infant formula [39]. These findings might be explained by the already established scientific fact that possessing sufficient knowledge about the risks and benefits of using a certain product can shape one’s attitude toward it as well as the decision to utilize the product. In this case, mothers who were well informed about the dangers of infant formula might have developed negative feelings toward the practice. This might have deterred the mothers in Debre Berhan city from providing infant formula to their babies.

In addition, our study revealed that mothers aged 25-34 years and 35-45 years were approximately three and five times more likely to feed infant formula, respectively, than were those aged 15-24 years. A similar finding was presented by a study conducted in Bahir Dar city, where mothers aged 52-34 years were found to have two and a half times greater odds of providing infant formula than were mothers aged 15-24 years [42]. In both instances, older maternal age was cited as one of the predisposing factors for infant formula feeding, possibly because younger mothers are less likely to have a stable job, a steady source of income, or spouses who can afford the high expenses associated with infant formula feeding. These situations might have prevented younger mothers from providing infant formula to their babies.

Another predictor of infant formula feeding identified by our study is parity. Those mothers who were primiparous were four and a half times more likely to practice infant formula feeding than were those who were multiparous. Similarly, a study in Mettu town showed that primiparous mothers were two times more likely to practice infant formula feeding than their counterparts [39]. This finding might be viewed from multiple standpoints. First, from an experience standpoint, prime mothers might have lacked the adequate knowledge and skills required for successful breastfeeding, and without proper breastfeeding support, they are more likely to face breastfeeding-related problems, including issues with proper breast attachment. As a result, they might prefer the much simpler method of child feeding, infant formula feeding, over complex and demanding breastfeeding. Second, from an economic standpoint, it is much more difficult for multiparous mothers to feed one child infant formula while taking care of their other children’s needs.

Unfortunately, our study also revealed that mothers who had received ANC were approximately two times more likely to practice infant formula feeding than were those who had not received ANC. However, given that a link between healthcare providers and breast milk substitute manufacturing companies has been found, this finding may not be surprising. One of the aggressive promotional tactics used by these companies is the use of healthcare providers as salespeople. These companies lobby healthcare providers to offer free samples and unnecessarily push mothers to use infant formula as an alternative food for their babies during ANC visits [9]. However, such tactics for promoting breast milk substitutes strongly contradict the WHO international code of marketing breast milk substitutes and jeopardize the trust between mothers and healthcare providers.

Finally, the odds of infant formula feeding were approximately four times greater among mothers who had delivered via C/S than among those who had not. Similarly, studies from Addis Ababa city, Dire Dawa city, and Egypt have shown that the odds of receiving infant formula are greater among mothers who underwent C/S delivery [41,44,47]. This finding might arise from the fact that mothers who undergo C/S delivery experience significant postoperative pain and fatigue due to the trauma inflicted on their bodies, and coupled with the poorly established pain management system in developing countries, in the postoperative period, mothers experience significant pain and are unable to perform skin-to-skin contact and breastfeed their babies promptly. The other reason might also be the separation of babies from their mothers following an operation, which can prevent mothers from being near their babies and breastfeeding them as needed.

## Conclusion

In Debre Berhan city, a significant number of mothers practiced infant formula feeding, and maternal age, number of parties, ANC utilization, mode of delivery, awareness of the risk of infant formula feeding, and breastfeeding initiation time were important predictors of infant formula feeding among the mothers living in the city. These findings imply that the majority of the factors that push mothers to practice infant formula feeding are modifiable if interventions focused on improving mothers’ and healthcare providers’ knowledge of the risk of infant formula feeding, the benefits of SVD, and the importance of optimal breastfeeding are designed and implemented. In addition, significant attention must also be directed toward breast milk substitute-producing companies, distributors, and healthcare providers to ensure that they abide by the rules and regulations that govern the promotion, sales, and recommendation of breast milk substitutes in Ethiopia and worldwide.

## Declarations

### Author Contributions

F.Z. and A.G. conceptualized the research idea, designed the study methodology, collected and analyzed the data, wrote the original manuscript draft, and reviewed and revised the manuscript; A.D. designed the study methodology, provided academic and research mentorship, contributed to the research design and methodology, reviewed and provided critical feedback on the manuscript, and approved the final version of the manuscript; M.A. designed the study methodology, provided academic and research mentorship, contributed to the research design and methodology, reviewed and provided critical feedback on the manuscript, and approved the final version of the manuscript; S.H. provided guidance and supervision throughout the research process, assisted in study design and methodology development, and reviewed and edited the manuscript for critical intellectual content.

### Institutional Review Board Statement

Ethical clearance was obtained from the ethical review board of the Asrat Woldeyes Health Science campus, Debre Berhan University, in 2023 (**protocol number: IRB-147**). Additionally, the city health office wrote a letter of cooperation, which was subsequently submitted to the responsible bodies. Furthermore, both verbal and written informed consent were obtained from each participant before each interview. To maintain the anonymity of the study participants, all their personal information was kept confidential.

### Consent to participate

Both verbal and written informed consent were obtained from the participants before each interview. To maintain the anonymity of the study participants, all their personal information was also kept confidential.

### Data availability statement

The datasets generated and/or analyzed during the current study are available from the corresponding author upon reasonable request.

### Consent for publication

Not applicable (NA)

## Supporting information

Questionnar

