## Supplementary material for "Determinants of infant formula feeding in Debre Berhan city: A community-based cross-sectional study": Questionnar

### Participant Consent form

Hello! My name is …………… I am here on behalf of, the investigator, which are employee of Debre Berhan University, Asrat Woldeyes Health Science Campus, and Department of Public Health. They are conducting research to assess the determinants of infant formula feeding among mothers of infant aged 0-6 months in Debre Berhan city, Ethiopia.

I am going to ask you questions related to infant formula feeding and associated factors. The information you provide will be kept confidential and used only for this study. Only the investigators will have access to this information. If you do not want to answer any of the questions, you have the right to do so. However, if you are willingness to answer all of questions it is much appreciated.

Do you give consent to participate in this study? -----

Yes --------	No---------

Name and Signature of study participants’ ______________ Date _________

Name and signature of data collector ____________________

**Instruction**: This questionnaire was designed for face-to-face interview with mothers of infants aged 0-6 months.

**Note**: This questionnaire must be filled only by the interviewer once informed consent is obtained from the respondents. Place the answer in blank spaces for open ended questions and circles for multiple choice responses.

**Part 1: Sociodemographic characteristics**

| S.no | Questions | Answer | Skip |
| --- | --- | --- | --- |
| 1 | Maternal age | -------------years |  |
| 2 | What is your current marital status? | 1.Married  2. Single  3.Divorced  4.Widowed |  |
| 3 | What is your level of education? | 1.Unable to read and write  2. Able to read and write  3. Primary education  4. Secondary education  5. College and above |  |
| 4 | What is your religion? | 1. Orthodox  2. Protestant  3. Muslim  4. Other |  |
| 5 | Where do you reside? | 1.Urban  2. Rural |  |
| 6 | What is your current occupation? | 1.Private employee  2.Goverment employee  3.Daily laborer  4.Trader/Private company  5.Farmer  6. Housewife  7. Other |  |
| 7 | What is your husband level of education? | 1.Unable to read and write  2. Able to read and write  3. Primary education  4. Secondary education  5. College and above |  |
| 8 | What is your husband’s current occupation? | 1.Private employee  2.Goverment employee  3.Daily laborer  4.Trader/Private company  5.Farmer  6. Other (Specify)------- |  |
| 9 | Age of your current child | -----------------months |  |
| 10 | Age difference between your last consecutive children? | -----------------years |  |
| 11 | Sex of your current child? | 1. Male 2. Female |  |
| **Section III. Obstetrics and related health service questions.** | | | |
| 12 | Number of babies delivered in the last pregnancy | 1.single  2.twins  3. triplets |  |
| 13 | Estimated weight of your last child at birth? | ------------------Kg |  |
| 14 | How many children did you have? | ------------- |  |
| 15 | Did you receive ANC during your last child pregnancy? | 1.Yes  2.No |  |
| 16 | How many times did you receive ANC? | 1.Once  2. Two times  3.Three times  4.Four times and above |  |
| 17 | If yes, where did you receive ANC? | 1.Gov.t Health Facilities  2.Private Health facilities  3.other---------------- |  |
| 18 | Did you receive counseling about breast feeding during any of your ANC visit? | 1.Yes.  2.No |  |
| 19 | Where did you give birth to your current child? | 1.Gov.t Health Facilities  2.Private Health Facilities  3.Home  4. Other, specify---------- |  |
| 20 | What was the mode of delivery? | 1.Normal/vaginal  2.C/S  3. Instrumental |  |
| 21 | Did you receive PNC service? | 1.Yes  2.No |  |
| 22 | How soon after birth did you put your child to your breast? | 1.With in 1hr of birth  2.After 1 hour |  |
| 23 | Have you ever heard/seen any information about infant formula? | 1.Yes  2.No |  |
| 24 | If yes, from where? | 1.From radio/TV  2.From health professionals  3.From supermarket keepers  4.From friends/family  5.Other, specify--------------- |  |
| 25 | Did your current child receive infant formula within the past 24h? | 1.Yes  2.No |  |
| 26 | What was your reason to start formula feeding? | 1.Due to insufficient breast milk  2.Maternl illness  3.Child illness  4. didn’t have no enough time to breast feed  5.Formula milk is as good as breast milk  6.Formula milk is better than breast milk  7.Others------------------------- |  |
| 27 | Would you recommend infant formula to others? | 1.Yes  2.No |  |
| 28 | Do you know the risks of infant formula feeding? | 1.Yes  2.No |  |
| 29 | If yes, what are they? | ------------------ |  |
| 30 | Do you believe maternity leave is enough? | 1.Yes  2.No |  |
| 31 | Did your family /friends push you to use infant formula? | 1.Yes  2.No |  |
| **Knowledge Questions** | | | |
| 1 | Breast milk is adequate for babies in the 1^st^ 6 months | 1.Yes  2.No  3. I do not know |  |
| 2 | Breast milk protects children from childhood illnesses | 1.Yes  2.No  3. I do not know |  |
| 3 | Formula feeding is more costly than breast milk | 1.Yes  2.No  3. I do not know |  |
| 4 | Breast milk nutritious | 1.Yes  2.No  3. I do not know |  |
| 5 | Breastfeeding increases mother to child bond | 1.Yes  2.No  3. I do not know |  |
| 6 | Breastfeeding can prevent pregnancy | 1.Yes  2.No  3. I do not know |  |
| 7 | Breastfeeding has more advantage than formula feeding | 1.Yes  2.No  3. I do not know |  |
